# Assessment of mesalamine in the alterations of metabolic parameters in comorbid ulcerative colitis and metabolic syndrome patients: A retrospective study

**DOI:** 10.1101/2021.05.27.21257690

**Authors:** Graziella R. Paniz, Fray M. Arroyo-Mercado, Christina L. Ling, E. Eunice Choi, Harry E. Snow, Neal E. Rakov, Eliseo F. Castillo

## Abstract

**BACKGROUND & AIMS:** It is unclear how the gut targeting medication mesalamine alters metabolic parameters associated with Metabolic Syndrome (MetS). We completed a retrospective analysis on ulcerative colitis (UC) and MetS comorbid patients receiving mesalamine to examine the effects of mesalamine on the metabolic risk factors associated with MetS.

**METHODS:** We performed a retrospective chart review using Cerner’s Health Facts (from July 2007 to July 2017). We identified UC patients with a MetS comorbidity and who were prescribed mesalamine within +/- 7 days of an encounter in which they were diagnosed with UC. We then collected the patient’s blood pressure, labs, and body measurement index (BMI) for each of these patient at the index date and the closest values to 12 months after the index date. We used analysis of covariance (ANCOVA) to determine the effect of mesalamine therapy in patients with both UC and MetS on the metabolic parameters after 12 months of treatment compared to baseline.

**RESULTS:** Our search of Cerner Health Facts identified 6,197 UC patients with concomitant MetS who were prescribed mesalamine. Of these individuals, 48% were female and 52% were male and within this cohort 88.3% received oral mesalamine and 11.7% received mesalamine via the rectal route. Oral mesalamine reduced fasting glucose levels and increased HDL cholesterol in these patients. C-reactive protein levels and erythrocyte sedimentation rate were also significantly reduced. Rectal mesalamine only reduced BMI. Further analysis revealed several MetS conditions risk factors were further improved when mesalamine was taken in the absence of medication for hypertension, hyperglycemia or dyslipidemia.

**CONCLUSION:** In a retrospective chart study of UC-MetS patients, we found oral mesalamine improved several metabolic parameters associated with MetS. Our findings suggest the PPAR agonist mesalamine that targets the gastrointestinal tract could prove beneficial in improving hypertension, hyperglycemia and dyslipidemia.

## INTRODUCTIONS

Metabolic Syndrome (MetS) identifies a group of risk factors that increases an individual’s risk for several clinical conditions such as cardiovascular disease, stroke and cancer^1-10^. An individual must have at least three of the following risk factors to be diagnosed with MetS: abdominal (central) obesity, high blood pressure/hypertension, elevated blood sugar/insulin resistance (*i*.*e*., hyperglycemia), increased triglycerides and low high-density lipoprotein (HDL) cholesterol (*i*.*e*., dyslipidemia)^11^. Both hyperglycemia and dyslipidemia are also highly associated with non-alcoholic fatty liver disease (NAFLD) which may be the liver manifestation of MetS^12-20^. It is estimated over a quarter of the world’s population and up to one-third of United States adults have MetS^21-23^. Unfortunately, there are no medications approved to treat MetS nor NAFLD. Furthermore, the prescribed medications only control the risk factors associated with MetS including blood pressure, hyperglycemia and dyslipidemia. Given the current incidence and prevalence of MetS, it is critical to identify the etiological factors leading to this disease to provide targeted interventional strategies, as changes in lifestyle alone may not be enough to reverse these conditions.

Recent studies in humans and rodents have suggested a role for the gastrointestinal (GI) tract in contributing to numerous diseases and disorders associated with metabolic dysfunction such as dyslipidemia and hyperglycemia^24-38^. Currently, peroxisome proliferator–activated receptor (PPAR) agonist are therapeutically targeted to treat dyslipidemia and hyperglycemia ^39-50^. PPARs are a group of transcription factors involved in numerous aspects of cellular differentiation and function including the regulation of energy homeostasis and inflammation ^51-57^. In the GI tract, PPAR-γ activity is decreased in individuals diagnosed with inflammatory bowel disease (IBD) such as ulcerative colitis (UC) and Crohn’s disease^58-62^. Interestingly, there are some individuals who present with UC and have comorbid conditions that are linked with MetS such as dyslipidemia, insulin resistance, hypertension, and increased abdominal obesity^63-69^. NAFLD is also frequently reported in patients with IBD^63, 70, 71^. Mesalamine, a PPAR agonist, has been documented to have beneficial effects in active, mild to moderate UC^72-74^. It has also been used successfully in maintenance therapy for UC^75^.

Mesalamine is an aminosalicylate compound also known as 5-aminosalicylic acid (5-ASA). Mesalamine is only approved for IBD but is considered relatively safe for individuals making it a great therapeutic drug to repurpose for MetS and NAFLD. Therefore, we performed a retrospective analysis on individuals who were diagnosed with UC, received mesalamine, and had at least three of the five following MetS conditions^13, 76-78^: hypertriglyceridemia, low HDL, hypertension, hyperglycemia, and a BMI over 27 to determine the beneficial effects of targeting the GI tract with a PPAR agonist on these metabolic parameters.

## METHODS

### Data Acquisition

All data were collected according to our institutional review board-approved protocol (20-369). Our study was a retrospective chart review, involving the use of existing data with no or minimal risk to participants, it had an institutional review board and patient consent exempt status under human subject regulations. All data used in this study come from Cerner Health Facts database. There were 478,236,643 unique encounters from 68,696,329 unique patients in Health Facts. Data in Health Facts was extracted directly from the electronic medical records (EMR) from hospitals in which Cerner has a data use agreement. Encounters included pharmacy, clinical and microbiology laboratory, admission, and billing information from affiliated patient care locations. All admissions, medication orders and dispensing, laboratory orders and specimens were date and time stamped, providing a temporal relationship between treatment patterns and clinical information. Cerner Corporation has established Health Insurance Portability and Accountability Act-compliant operating policies to establish de-identification for Health Facts. In all, Health Facts was used to find individuals who were diagnosed with UC with a MetS comorbidity that received mesalamine.

### Data Inclusion

We found patients between July 1, 2007 and July 1, 2017 who had International Classification of Diseases (ICD) code for UC. All patients were age 18 or older. This yielded 52,676 patients. Among these patients we found patients who also had a mesalamine prescription within +/- 7 days of an encounter in which they were diagnosed with UC. Mesalamine included sulfasalazine and 5-aminosalycyclic acid. For each patient with UC who also had a mesalamine prescription, we identified the mesalamine start date as the index date. From this, we then found all relevant diagnoses, labs, medications of interest, blood pressures, body mass index (BMI) measurements, heights and weights in the medical record within +/- 12 months of each patient’s index date. This information was used to calculate the five risk factors of metabolic syndrome. This included increased blood pressure (130/85 mmHg), high blood sugar levels (≥110 mg/dL), high triglycerides (≥150 mg/dL), low levels of HDL (Men <40mg/dL; Women <50mg/dL), and overweight status as defined by BMI (≥ 25.0 kg/m^2^). In computing factors, a diagnosis of obesity was enough to satisfy the factor of BMI >27. Height and weight were used to compute BMI when BMI was not available. There were 6,197 patients who had UC with a MetS comorbidity who were prescribed mesalamine within +/- 7 days of an encounter in which they were diagnosed with UC. For each of these patient, we looked for the closest values before the index date, and then the closest values to 12 months after the index date for the patient’s blood pressure, labs, and BMI.

### Statistical Analysis

Paired t-test or Wilcoxon Rank Sum test were used for continuous variables, and McNemar’s test for categorical variables. For inferential statistics, we used analysis of covariance (ANCOVA) to determine the effect of mesalamine therapy in patients with both UC and MetS on the metabolic parameters after 12 months of treatment compared to baseline.

## RESULTS

### Identified Population and Use of Mesalamine

Our search of Cerner Health Facts identified 52,676 UC patients between July 2007 and July 2017. Of these individuals, 6,197 UC patients with concomitant MetS and were prescribed mesalamine within +/- 7 days of an encounter in which they were diagnosed with UC. MetS complications included diabetes (19.7%), hypertension (45.6%), and hyperlipidemia (9.7%) (**Table 1**). Approximately 48% of the individuals identified were female and 52% were male with an overall mean age of 53.8 (±19.9) (**Table 1**). Within this cohort, 88.3% received mesalamine orally and 11.7% via the rectal route (**Table 1**).

**Table 1.** Demographics of Cohort, Mesalamine usage and Metabolic Syndrome complications among the Cohort.

| Characteristics | Total patients<br>n=6197 |
| --- | --- |
| Age, mean ± SD | 53.8 ± 19.9 |
| Sex: |  |
| Female | 3275 (52.9%) |
| Male | 2922 (47.2%) |
| Race: |  |
| White | 4836 (78.0%) |
| Black | 814 (13.1%) |
| Asian | 62 (1.0%) |
| Native American | 59 (1.0%) |
| Hispanic | 46 (0.7%) |
| Other | 380 (6.1%) |
| Product strength: |  |
| 250 mg | 542 (10.4%) |
| 375 mg | 80 (1.5%) |
| 400 mg | 2520 (48.5%) |
| 500 mg | 928 (17.9%) |
| 1000 mg | 388 (7.5%) |
| 1200 mg | 735 (14.2%) |
| Oral | 5472 (88.3%) |
| Rectal | 725 (11.7%) |
| Dose Form: |  |
| Capsule, extended release | 756 (12.2%) |
| Delayed release capsule | 1030 (16.6%) |
| Enema | 329 (5.3%) |
| Enteric coated tablet | 2999 (48.4%) |
| Suppository | 396 (6.4%) |
| Tablet | 686 (11.1%) |
| Metabolic Syndrome complications: |  |
| Diabetes | 1220 (19.7%) |
| Hypertension | 2825 (45.6%) |
| Hyperlipidemia | 598 (9.7%) |

### Influence of oral mesalamine on metabolic parameters in UC-MetS comorbid patients

For the 5,472 individuals taking oral mesalamine that had an ICD code for both UC and MetS risk factors, we examined specific metabolic parameters (HDL cholesterol, triglycerides, fasting glucose, blood pressure, and BMI) in these individuals at baseline and after 12 months on mesalamine. We found the levels of HDL cholesterol were significantly increased (P < 0.001) and fasting glucose levels were significantly decreased (P < 0.001) (**Table 2**) after 12 months on oral mesalamine. No significant change was observed for BMI, triglyceride levels, or systolic blood pressure; however, diastolic blood pressure was raised from 70.1 mmHg to 71.6 mmHg (P < 0.001) on mesalamine (**Table 2**). Oral mesalamine usage also significantly reduced C-reactive protein (CRP) levels (P < 0.001) as well as the erythrocyte sedimentation rate (ESR) (P < 0.001) in UC-MetS comorbid patients (**Table 2**). Comparing the different doses of oral mesalamine on HDL cholesterol levels, fasting glucose, CRP and ESR (**Supplementary Table 1**), we found high dose (1200 – 1000 mg) mesalamine had an average increase in HDL cholesterol levels of 48.9 mg/dL while medium dose (500, 400, 375 mg) was 43.8 mg/dL, and low dose (250 mg) was 42.7 mg/dL. Average fasting glucose levels were 142.0 mg/dL for high dose mesalamine, 139.3 mg/dL for medium dose, and 142.0 mg/dL for low dose (**Supplementary Table 1**). For CRP, high dose mesalamine had the lowest CRP average at 3.9 mg/dL, while medium dose was 4.6 mg/dL and low dose was 6.1 mg/dL. ESR followed the same trend with high dose mesalamine having an average ESR at 41.8 mm/h, while medium dose was 42.2 mm/h and low dose was 44.8 mm/h (**Supplementary Table 1**).

**Table 2.** Comparison of metabolic parameters in 5,472 UC-MetS Patients taking oral mesalamine. BUN, blood urea nitrogen; HDL, high-density lipoprotein; LDL, low-density lipoprotein.

| Measurement | Baseline | 12 months | difference | Lower 95% | Upper 95% | p value |
| --- | --- | --- | --- | --- | --- | --- |
| Body Mass Index (BMI) | 31.2 | 31.0 | -0.2 | -0.4 | 0.1 | 0.17 |
| Systolic Blood pressure (mmHg) | 127.4 | 128.3 | 0.8 | -0.2 | 1.8 | 0.10 |
| Diastolic Blood pressure (mmHg) | 70.1 | 71.6 | 1.5 | 0.8 | 2.1 | <0.01* |
| Total Cholesterol (mg/dL) | 159.9 | 159.9 | 0.1 | -2.7 | 2.9 | 0.96 |
| HDL cholesterol (mg/dL) | 44.3 | 46.0 | 1.7 | 0.8 | 2.5 | <0.01* |
| LDL cholesterol (mg/dL) | 91.7 | 90.2 | -1.5 | -4.0 | 1.1 | 0.25 |
| Triglycerides (mg/dL) | 129.4 | 130.4 | 1.0 | -4.1 | 6.2 | 0.95 |
| Fasting glucose (mg/dL) | 142.7 | 139.5 | -3.2 | -5.2 | -1.2 | <0.01* |
| BUN (mg/dL) | 18.2 | 19.5 | 1.2 | 0.8 | 1.6 | <0.01* |
| C-Reactive Protein (mg/dL) | 5.5 | 4.2 | -1.3 | -1.7 | -0.8 | <0.01* |
| Alkaline Phosphatase (IU/L) | 89.0 | 93.3 | 4.3 | 1.2 | 7.4 | <0.01* |
| Albumin (g/L) | 3.1 | 3.1 | 0.1 | 0.0 | 0.1 | 0.01* |
| Erythrocyte Sedimentation Rate (mm/h) | 44.3 | 41.5 | -2.7 | -4.5 | -1.0 | <0.01* |

Given some UC-MetS comorbid patients were on medication (**Supplementary Table 2**) to treat hypertension, diabetes or hyperlipidemia, we assessed if the effects described above were dependent on these other medications. Surprisingly, we found individuals on oral mesalamine without hypertension medication had a significant decrease in systolic and diastolic blood pressure (P < 0.01) as well as BMI (P < 0.01) and fasting glucose levels (P < 0.01) compared to individuals taking both mesalamine and hypertension medication (**Supplementary Table 3**). Assessing individuals taking mesalamine without anti-diabetic medication, we observed a significant decrease in BMI (P < 0.01) and fasting glucose levels (P < 0.01) as well as an increase in HDL cholesterol levels (P < 0.05) than individuals taking both mesalamine and anti-diabetic medication (**Supplementary Table 4)**. When individuals were taking mesalamine and medications to treat hyperlipidemia (**Supplementary Table 5**), a significant decrease in total cholesterol (P = 0.01) and low-density lipoprotein (LDL) cholesterol levels (P = 0.01) were observed. Significant changes were also observed for both systolic and diastolic blood pressure. However, triglyceride (P = 0.01) and fasting glucose (P < 0.01) levels were significantly lower in individuals taking mesalamine without statins or cholesterol lowering medication. (**Supplementary Table 5**). No major difference were observed for CRP or ESR when individuals were on mesalamine with or without medications to treat hypertension, diabetes or hyperlipidemia (**Supplementary Table 3-5**).

### Influence of rectal mesalamine on metabolic parameters in UC-MetS comorbid patients

Cerner Health Facts identified 725 individuals taking mesalamine via the rectal route. Analysis of the various metabolic parameters (*i*.*e*., HDL cholesterol, triglycerides, fasting glucose, blood pressure, and BMI) in these UC-MetS comorbid patients at baseline and after 12 months on rectal mesalamine, we only found a statistically significant reduction in BMI after mesalamine treatment (**Table 3**). Additionally, the average levels of CRP and ESR were reduced; however, these changes were not significant (**Table 3**). These parameters were also assessed for individuals taking medication for hypertension, diabetes or hyperlipidemia while on rectal mesalamine. Once again individuals taking rectal mesalamine in the absence of medication for hypertension had a significant reduction in systolic blood pressure (P < 0.01), BMI (P = 0.04), and fasting glucose levels (P = 0.01) (**Supplementary Table 6**). Fasting glucose levels were also significantly reduced in individuals taking rectal mesalamine in the absence of anti-diabetic medication (P < 0.01) (**Supplementary Table 7**). No major difference were observed for individuals taking rectal mesalamine and medication to treat hyperlipidemia (**Supplementary Table 8**). No major difference were observed for CRP or ESR when individuals were on rectal mesalamine with or without medications to treat hypertension, diabetes or hyperlipidemia (**Supplementary Table 6-8**). Lastly, alkaline phosphatase levels were significantly reduced when individuals were receiving rectal mesalamine in the absence of medication for diabetes or dyslipidemia (**Supplementary Table 7-8**).

**Table 3.** Comparison of metabolic parameters in 725 UC-MetS Patients taking rectal mesalamine. BUN, blood urea nitrogen; HDL, high-density lipoprotein; LDL, low-density lipoprotein.

| Measurement | Baseline | 12 months | difference | Lower 95% | Upper 95% | p value |
| --- | --- | --- | --- | --- | --- | --- |
| Body Mass Index (BMI) | 27.9 | 27.1 | -0.8 | -1.6 | 0.0 | 0.02* |
| Systolic Blood pressure (mmHg) | 138.2 | 138.2 | 0.0 | -2.8 | 2.9 | 0.97 |
| Diastolic Blood pressure (mmHg) | 75.5 | 76.2 | 0.8 | -1.1 | 2.6 | 0.42 |
| Total Cholesterol (mg/dL) | 133.9 | 139.8 | 5.9 | -9.6 | 21.4 | 0.45 |
| HDL cholesterol (mg/dL) | 43 | 44.7 | 1.7 | -3.5 | 6.9 | 0.52 |
| LDL cholesterol (mg/dL) | 82.3 | 76.4 | -5.9 | -19.8 | 8.0 | 0.40 |
| Triglycerides (mg/dL) | 97.7 | 75.7 | -22.1 | -41.8 | -2.4 | 0.11 |
| Fasting glucose (mg/dL) | 162.1 | 161.1 | -1.0 | -7.6 | 5.6 | 0.43 |
| BUN (mg/dL) | 20.8 | 22.8 | 2.0 | 0.6 | 3.4 | 0.01* |
| C-Reactive Protein (mg/dL) | 10.8 | 9.7 | -1.1 | -2.2 | 0.1 | 0.06 |
| Alkaline Phosphatase (IU/L) | 133.7 | 139.1 | 5.4 | -2.2 | 12.9 | 0.19 |
| Albumin (g/L) | 2.6 | 2.7 | 0.1 | -0.1 | 0.2 | 0.34 |
| Erythrocyte Sedimentation Rate (mm/h) | 31.0 | 26.8 | -4.2 | -11.7 | 3.3 | 0.30 |

## DISCUSSION

In this retrospective study, we examined the beneficial effects of mesalamine on various metabolic parameters associated with MetS. Given mesalamine is only approved to treat mild to moderate UC, the effects of mesalamine were examined in UC patients who also had MetS. Using Cerner Health Facts, we identified UC-MetS comorbid patients receiving both oral and rectal mesalamine. In our assessment, we found oral mesalamine proved beneficial in improving several metabolic parameters. This included increasing HDL cholesterol levels and reducing fasting glucose levels (**Table 2**). Other factors that are frequently increased in MetS such as CRP levels and increased ESR but also associated with UC were also significantly decreased^79-83^. Interestingly, oral mesalamine proved beneficial for other metabolic parameters (BMI, blood pressure, triglycerides) when individuals were not on medications to treat hypertension, diabetes or dyslipidemia (**Supplementary Table 3-5**). Rectal mesalamine was associated with decreased BMI and appeared more impactful when taken in the absence of medications for hypertension. Overall, our findings suggest targeting the gut with the PPAR agonist mesalamine could prove beneficial in improving metabolic parameters associated with MetS.

While our study has several limitations including underlying chronic intestinal inflammation, lack of data for steroid use, and reliance on documentation and coding, it is the first large retrospective study that considers the effect of mesalamine on metabolic parameters associated with MetS. We suspect the true effect of mesalamine on these MetS risk factors may be underrepresented due to the chronic intestinal inflammation that characterizes UC. Nevertheless, mesalamine has proved beneficial in reducing fasting glucose levels and hepatic steatosis in an animal model of diet-induced obesity^84^. Within the last decade, numerous studies have shown the GI tract contributes to metabolic dysfunction^24-38^. Specifically, chronic consumption of a high-fat diet (HFD) enhances intestinal barrier defects leading to enhanced intestinal permeability (IP), dysbiosis (i.e. alteration/reduction in microbial diversity and metabolites), and low-level of inflammation^38, 85-89^. Dysbiosis can also increase IP as well as modulate the intestinal immune system^90-92^. Both IP and dysbiosis can result in low-grade inflammation that affects weight gain, hyperglycemia and insulin resistance, dyslipidemia, and hypertension^28-30, 93-95^. In addition, increased IP and dysbiosis have been observed in individuals who have been diagnosed with NAFLD (as well as individuals who develop nonalcoholic steatohepatitis, NASH), type 2 diabetes, and obesity ^25-27, 93, 96-106^. The mechanism of action of mesalamine is believed to work on three areas of the gut to maintain intestinal homeostasis: the microbiota, intestinal barrier, and the immune system^84, 107-116^. Mesalamine can directly inhibit proinflammatory pathways NF-κB and MAPK that promote the expression of proinflammatory cytokines (*e.g*., TNFα, IL-6 and IL-1β)^84, 117-123^. Mesalamine also dampens COX-2 expression that leads to PGE2 and PGF2 production^117, 120, 123-126^. Recent reports have demonstrated mesalamine increases the diversity of the gut microbiota through metabolic changes and direct effects on the microbiota^113, 127-129^. Lastly, mesalamine has been shown to decrease IP and restore epithelial barrier function^84, 111, 112^. Therefore, targeting the gut with mesalamine in individuals with MetS or NAFLD could prove beneficial in treating metabolic disorders.

## Data Availability

The datasets generated and analyzed during this study are not publicly available because of institutional review board restrictions, but data acquisition is described in methods.

## ACKNOWLEDGMENTS

Supported in part by the National Center for Research Resources and the National Center for Advancing Translational Sciences of the National Institutes of Health (NIH) through grant no. UL1TR001449 (E.F.C.) and NIH Clinical and Translational Science Award to UNMHSC, the Biostatistics, Epidemiology, and Research Design (BERD) Core of the UNM HSC Clinical & Translational Science Center (CTSC) through grant no. UL1TR001449. C.L.L is now part of the Division of Gastroenterology, Department of Medicine, University of California, Irvine, Irvine, CA 92697.

## AUTHOR CONTRIBUTIONS

G.R.P., C.L.L. and E.F.C. designed the study. H.S. and E.H.C. contributed to data acquisition and statistical analysis. G.R.P, F.M.A-M, C.L.L., N.E.R. and E.F.C. analyzed and interpreted data. G.R.P., F.M.A-M, and E.F.C wrote the first draft of the manuscript. All authors reviewed, edited and approved the manuscript.

## DATA AVAILABILITY

**Supplementary Table 1.** Dose effect of oral mesalamine on metabolic parameters in 5,472 UC-MetS Patients.

| **Oral mesalamine (dose)** | **high** | **med** | **low** |
| --- | --- | --- | --- |
| Body Mass Index (BMI) | 31.5 | 31.3 | 30.6 |
| Systolic Blood pressure (mmHg) | 129.3 | 128.0 | 126.4 |
| Diastolic Blood pressure (mmHg) | 72.4 | 70.5 | 69.6 |
| Cholesterol (mg/dL) | 160.4 | 153.7 | 165.6 |
| HDL cholesterol (mg/dL) | 48.9 | 43.8 | 42.7 |
| LDL cholesterol (mg/dL) | 89.3 | 86.8 | 96.8 |
| Triglycerides (mg/dL) | 139.1 | 127.8 | 122.8 |
| Fasting glucose (mg/dL) | 142.0 | 139.3 | 142.0 |
| BUN (mg/dL) | 18.8 | 18.8 | 18.9 |
| C-Reactive Protein (mg/dL) | 3.9 | 4.6 | 6.1 |
| Alkaline Phosphatase (IU/L) | 87.8 | 92.3 | 93.4 |
| Albumin (g/L) | 3.1 | 3.2 | 3.1 |
| Erythrocyte Sedimentation Rate (mm/h) | 41.8 | 42.2 | 44.8 |

**Supplementary Table 2.**
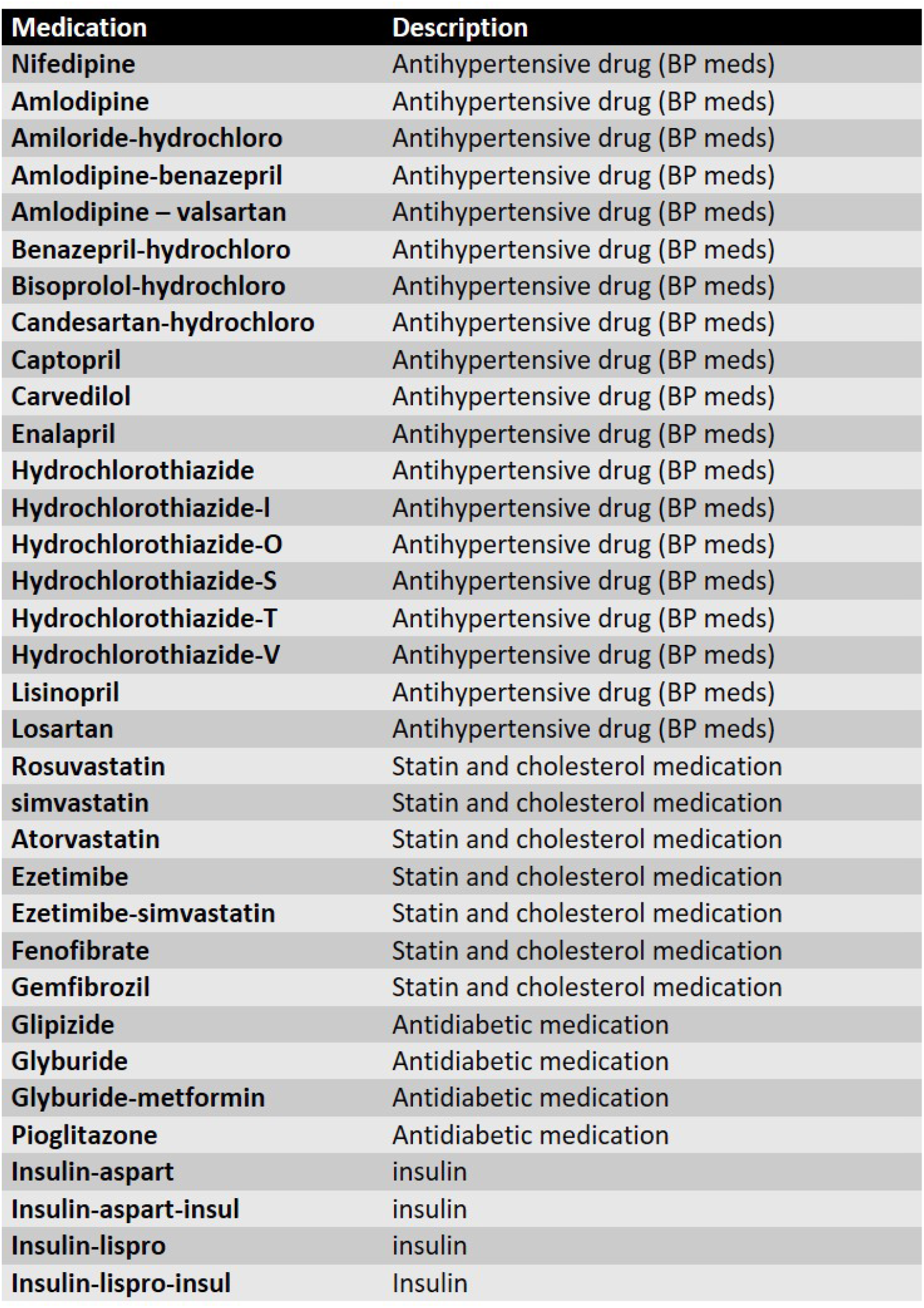
List of medications to treat hypertension, dyslipidemia or hyperglycemia in study cohort.

**Supplementary Table 3.** Comparison of metabolic parameters in UC-MetS Patients taking oral mesalamine with or without blood pressure medication. BP, blood pressure medication.

| **Oral mesalamine** | **Without BP Meds** | **With BP Meds** | **P value** |
| --- | --- | --- | --- |
| Body Mass Index (BMI) | 30.3 | 32.0 | <0.01* |
| Systolic Blood pressure (mmHg) | 122.0 | 133.7 | <0.01* |
| Diastolic Blood pressure (mmHg) | 69.5 | 72.2 | <0.01* |
| Total Cholesterol (mg/dL) | 162.6 | 157.1 | 0.06 |
| HDL cholesterol (mg/dL) | 45.2 | 45.0 | 0.84 |
| LDL cholesterol (mg/dL) | 91.9 | 90.1 | 0.48 |
| Triglycerides (mg/dL) | 128.7 | 131.1 | 0.15 |
| Fasting glucose (mg/dL) | 137.3 | 144.9 | <0.01* |
| BUN (mg/dL) | 16.9 | 20.8 | <0.01* |
| C-Reactive Protein (mg/dL) | 5.0 | 4.8 | 0.67 |
| Alkaline Phosphatase (IU/L) | 94.3 | 88.0 | 0.41 |
| Albumin (g/L) | 3.1 | 3.1 | 0.55 |
| Erythrocyte Sedimentation Rate (mm/h) | 42.4 | 43.4 | 0.55 |

**Supplementary Table 4.** Comparison of metabolic parameters in UC-MetS Patients taking oral mesalamine with or without antidiabetic medication.

| **Oral mesalamine** | **Without Antidiabetic Meds** | **With Antidiabetic Meds** | **P value** |
| --- | --- | --- | --- |
| Body Mass Index (BMI) | 28.6 | 33.7 | <0.01* |
| Systolic Blood pressure (mmHg) | 127.2 | 128.6 | 0.57 |
| Diastolic Blood pressure (mmHg) | 71.5 | 70.1 | 0.35 |
| Cholesterol (mg/dL) | 164.1 | 155.7 | 0.22 |
| HDL cholesterol (mg/dL) | 47.5 | 42.8 | 0.05* |
| LDL cholesterol (mg/dL) | 92.9 | 89.0 | 0.48 |
| Triglycerides (mg/dL) | 130.1 | 129.7 | 0.59 |
| Fasting glucose (mg/dL) | 121.3 | 160.9 | <0.01* |
| BUN (mg/dL) | 17.2 | 20.5 | <0.01* |
| C-Reactive Protein (mg/dL) | 5.1 | 4.6 | 0.72 |
| Alkaline Phosphatase (IU/L) | 90.6 | 91.7 | 0.78 |
| Albumin (g/L) | 3.1 | 3.2 | 0.36 |
| Erythrocyte Sedimentation Rate (mm/h) | 38.0 | 47.8 | 0.07 |

**Supplementary Table 5.** Comparison of metabolic parameters in UC-MetS Patients taking oral mesalamine with or without medication to treat high cholesterol.

| **Oral mesalamine** | **Without Cholesterol Meds** | **With Cholesterol Meds** | **P value** |
| --- | --- | --- | --- |
| Body Mass Index (BMI) | 30.9 | 31.4 | 0.25 |
| Systolic Blood pressure (mmHg) | 126.6 | 129.2 | 0.01* |
| Diastolic Blood pressure (mmHg) | 71.9 | 69.8 | <0.01* |
| Cholesterol (mg/dL) | 163.6 | 156.1 | 0.01* |
| HDL cholesterol (mg/dL) | 45.5 | 44.8 | 0.55 |
| LDL cholesterol (mg/dL) | 94.6 | 87.4 | 0.01* |
| Triglycerides (mg/dL) | 126.4 | 133.4 | 0.01* |
| Fasting glucose (mg/dL) | 137.9 | 144.3 | <0.01* |
| BUN (mg/dL) | 17.0 | 20.7 | <0.01* |
| C-Reactive Protein (mg/dL) | 5.0 | 4.7 | 0.40 |
| Alkaline Phosphatase (IU/L) | 91.1 | 91.2 | 0.83 |
| Albumin (g/L) | 3.1 | 3.2 | 0.10 |
| Erythrocyte Sedimentation Rate (mm/h) | 42.1 | 43.7 | 0.24 |

**Supplementary Table 6.** Comparison of metabolic parameters in UC-MetS Patients taking rectal mesalamine with or without blood pressure medication. BP, blood pressure medication.

| **Rectal mesalamine** | **Without BP Meds** | **With BP Meds** | **P value** |
| --- | --- | --- | --- |
| Body Mass Index (BMI) | 25.9 | 29.2 | 0.04* |
| Systolic Blood pressure (mmHg) | 132.4 | 144.0 | <0.01* |
| Diastolic Blood pressure (mmHg) | 74.9 | 76.8 | 0.30 |
| Cholesterol (mg/dL) | 136.9 | 136.7 | 0.98 |
| HDL cholesterol (mg/dL) | 46.1 | 41.6 | 0.35 |
| LDL cholesterol (mg/dL) | 76.4 | 82.3 | 0.47 |
| Triglycerides (mg/dL) | 70.6 | 102.8 | 0.07 |
| Fasting glucose (mg/dL) | 153.5 | 169.8 | 0.01* |
| BUN (mg/dL) | 19.8 | 23.7 | <0.01* |
| C-Reactive Protein (mg/dL) | 10.0 | 10.6 | 0.69 |
| Alkaline Phosphatase (IU/L) | 144.5 | 128.3 | 0.52 |
| Albumin (g/L) | 2.8 | 2.5 | 0.09 |
| Erythrocyte Sedimentation Rate (mm/h) | 22.3 | 35.5 | 0.13 |

**Supplementary Table 7.** Comparison of metabolic parameters in UC-MetS Patients taking rectal mesalamine with or without antidiabetic medication.

| Rectal mesalamine | Without Antidiabetic Meds | With Antidiabetic Meds | P value |
| --- | --- | --- | --- |
| Body Mass Index (BMI) | 24.2 | 30.8 | 0.22 |
| Systolic Blood pressure (mmHg) | 134.2 | 142.2 | 0.37 |
| Diastolic Blood pressure (mmHg) | 77.1 | 74.6 | 0.64 |
| Cholesterol (mg/dL) | 160.4 | 113.3 | 0.17 |
| HDL cholesterol (mg/dL) | 47.0 | 40.6 | 0.65 |
| LDL cholesterol (mg/dL) | 108.5 | 50.1 | 0.11 |
| Triglycerides (mg/dL) | 108.9 | 64.4 | 0.60 |
| Fasting glucose (mg/dL) | 123.6 | 199.6 | <0.01* |
| BUN (mg/dL) | 15.7 | 27.8 | <0.01* |
| C-Reactive Protein (mg/dL) | 8.0 | 12.6 | 0.20 |
| Alkaline Phosphatase (IU/L) | 97.0 | 175.7 | 0.02* |
| Albumin (g/L) | 2.9 | 2.4 | 0.27 |
| Erythrocyte Sedimentation Rate (mm/h) | 26.8 | 31.0 | 0.61 |

**Supplementary Table 8.** Comparison of metabolic parameters in UC-MetS Patients taking rectal mesalamine with or without medication to treat high cholesterol.

| Rectal mesalamine | Without Cholesterol Meds | With Cholesterol Meds | P value |
| --- | --- | --- | --- |
| Body Mass Index (BMI) | 26.3 | 28.8 | 0.23 |
| Systolic Blood pressure (mmHg) | 136.7 | 139.7 | 0.43 |
| Diastolic Blood pressure (mmHg) | 76.8 | 74.9 | 0.41 |
| Cholesterol (mg/dL) | 136.4 | 137.2 | 0.94 |
| HDL cholesterol (mg/dL) | 46.1 | 41.6 | 0.37 |
| LDL cholesterol (mg/dL) | 76.5 | 82.2 | 0.50 |
| Triglycerides (mg/dL) | 82.3 | 91.1 | 0.67 |
| Fasting glucose (mg/dL) | 160.4 | 162.8 | 0.68 |
| BUN (mg/dL) | 20.4 | 23.1 | 0.05* |
| C-Reactive Protein (mg/dL) | 11.2 | 9.4 | 0.37 |
| Alkaline Phosphatase (IU/L) | 117.6 | 155.2 | 0.03* |
| Albumin (g/L) | 2.7 | 2.6 | 0.61 |
| Erythrocyte Sedimentation Rate (mm/h) | 32.6 | 25.2 | 0.31 |

## REFERENCES

1. Zhang J, Wu H, Wang R. Metabolic syndrome and esophageal cancer risk: a systematic review and metaanalysis. Diabetol Metab Syndr 2021;13:8.

2. Tune JD, Goodwill AG, Sassoon DJ, et al. Cardiovascular consequences of metabolic syndrome. Transl Res 2017;183:57–70.

3. Mariani M, Sassano M, Boccia S. Metabolic syndrome and gastric cancer risk: a systematic review and meta-analysis. Eur J Cancer Prev 2021;30:239–250.

4. Kwon HM, Kim BJ, Lee SH, et al. Metabolic syndrome as an independent risk factor of silent brain infarction in healthy people. Stroke 2006;37:466–70.

5. Han AL. Association of Cardiovascular Risk Factors and Metabolic Syndrome with non-alcoholic and alcoholic fatty liver disease: a retrospective analysis. BMC Endocr Disord 2021;21:91.

6. Gonzalez-Chavez A, Chavez-Fernandez JA, Elizondo-Argueta S, et al. Metabolic Syndrome and Cardiovascular Disease: A Health Challenge. Arch Med Res 2018;49:516–521.

7. Dong S, Wang Z, Shen K, et al. Metabolic Syndrome and Breast Cancer: Prevalence, Treatment Response, and Prognosis. Front Oncol 2021;11:629666.

8. DeBoer MD, Filipp SL, Sims M, et al. Risk of Ischemic Stroke Increases Over the Spectrum of Metabolic Syndrome Severity. Stroke 2020;51:2548–2552.

9. Chen H, Zheng X, Zong X, et al. Metabolic syndrome, metabolic comorbid conditions and risk of early-onset colorectal cancer. Gut 2021;70:1147–1154.

10. Chaiyasit K, Wiwanitkit V. Underlying metabolic syndrome, cancer and brain infarction: the story line. Diabetes Metab Syndr 2012;6:214.

11. Sherling DH, Perumareddi P, Hennekens CH. Metabolic Syndrome. J Cardiovasc Pharmacol Ther 2017;22:365–367.

12. Katsiki N, Perez-Martinez P, Anagnostis P, et al. Is Nonalcoholic Fatty Liver Disease Indeed the Hepatic Manifestation of Metabolic Syndrome? Curr Vasc Pharmacol 2018;16:219–227.

13. Yki-Jarvinen H. Non-alcoholic fatty liver disease as a cause and a consequence of metabolic syndrome. Lancet Diabetes Endocrinol 2014;2:901–10.

14. Rinaldi L, Pafundi PC, Galiero R, et al. Mechanisms of Non-Alcoholic Fatty Liver Disease in the Metabolic Syndrome. A Narrative Review. Antioxidants (Basel) 2021;10.

15. Lim S, Kim JW, Targher G. Links between metabolic syndrome and metabolic dysfunction-associated fatty liver disease. Trends Endocrinol Metab 2021.

16. Cho HJ, Hwang S, Park JI, et al. Improvement of Nonalcoholic Fatty Liver Disease Reduces the Risk of Type 2 Diabetes Mellitus. Gut Liver 2019;13:440–449.

17. Heo NY. Nonalcoholic Fatty Liver Disease Is a Stepping Stone in the Path toward Diabetes Mellitus. Gut Liver 2019;13:383–384.

18. Fujii H, Kawada N, Japan Study Group Of Nafld J-N. The Role of Insulin Resistance and Diabetes in Nonalcoholic Fatty Liver Disease. Int J Mol Sci 2020;21.

19. Cohen DE, Fisher EA. Lipoprotein metabolism, dyslipidemia, and nonalcoholic fatty liver disease. Semin Liver Dis 2013;33:380–8.

20. Amor AJ, Perea V. Dyslipidemia in nonalcoholic fatty liver disease. Curr Opin Endocrinol Diabetes Obes 2019;26:103–108.

21. Moore JX, Chaudhary N, Akinyemiju T. Metabolic Syndrome Prevalence by Race/Ethnicity and Sex in the United States, National Health and Nutrition Examination Survey, 1988-2012. Prev Chronic Dis 2017;14:E24.

22. Hirode G, Wong RJ. Trends in the Prevalence of Metabolic Syndrome in the United States, 2011-2016. JAMA 2020;323:2526–2528.

23. Saklayen MG. The Global Epidemic of the Metabolic Syndrome. Curr Hypertens Rep 2018;20:12.

24. Cani PD. Targeting gut microbiota with a complex mix of dietary fibers improves metabolic diseases. Kidney Int 2019;95:14–16.

25. Jiang W, Wu N, Wang X, et al. Dysbiosis gut microbiota associated with inflammation and impaired mucosal immune function in intestine of humans with non-alcoholic fatty liver disease. Sci Rep 2015;5:8096.

26. Gao R, Zhu C, Li H, et al. Dysbiosis Signatures of Gut Microbiota Along the Sequence from Healthy, Young Patients to Those with Overweight and Obesity. Obesity (Silver Spring) 2018;26:351–361.

27. Henao-Mejia J, Elinav E, Jin C, et al. Inflammasome-mediated dysbiosis regulates progression of NAFLD and obesity. Nature 2012;482:179–85.

28. Tilg H, Adolph TE. Influence of the human intestinal microbiome on obesity and metabolic dysfunction. Curr Opin Pediatr 2015;27:496–501.

29. Bleau C, Karelis AD, St-Pierre DH, et al. Crosstalk between intestinal microbiota, adipose tissue and skeletal muscle as an early event in systemic low-grade inflammation and the development of obesity and diabetes. Diabetes Metab Res Rev 2015;31:545–61.

30. Jouet P, Coffin B, Sabate JM. Small intestinal bacterial overgrowth in patients with morbid obesity. Dig Dis Sci 2011;56:615; author reply 615-6.

31. Stahel P, Xiao C, Nahmias A, et al. Role of the Gut in Diabetic Dyslipidemia. Front Endocrinol (Lausanne) 2020;11:116.

32. Rubino F, Forgione A, Cummings DE, et al. The mechanism of diabetes control after gastrointestinal bypass surgery reveals a role of the proximal small intestine in the pathophysiology of type 2 diabetes. Ann Surg 2006;244:741–9.

33. Tomkin GH. The intestine as a regulator of cholesterol homeostasis in diabetes. Atheroscler Suppl 2008;9:27–32.

34. Zhao M, Liao D, Zhao J. Diabetes-induced mechanophysiological changes in the small intestine and colon. World J Diabetes 2017;8:249–269.

35. Nascimento JC, Matheus VA, Oliveira RB, et al. High-Fat Diet Induces Disruption of the Tight Junction-Mediated Paracellular Barrier in the Proximal Small Intestine Before the Onset of Type 2 Diabetes and Endotoxemia. Dig Dis Sci 2020.

36. Ridaura VK, Faith JJ, Rey FE, et al. Gut microbiota from twins discordant for obesity modulate metabolism in mice. Science 2013;341:1241214.

37. Cani PD, Amar J, Iglesias MA, et al. Metabolic endotoxemia initiates obesity and insulin resistance. Diabetes 2007;56:1761–72.

38. Fayfman M, Flint K, Srinivasan S. Obesity, Motility, Diet, and Intestinal Microbiota-Connecting the Dots. Curr Gastroenterol Rep 2019;21:15.

39. Cariou B, Zair Y, Staels B, et al. Effects of the new dual PPAR alpha/delta agonist GFT505 on lipid and glucose homeostasis in abdominally obese patients with combined dyslipidemia or impaired glucose metabolism. Diabetes Care 2011;34:2008–14.

40. Patel H, Giri P, Patel P, et al. Preclinical evaluation of saroglitazar magnesium, a dual PPAR-alpha/gamma agonist for treatment of dyslipidemia and metabolic disorders. Xenobiotica 2018;48:1268–1277.

41. Jain N, Bhansali S, Kurpad AV, et al. Effect of a Dual PPAR alpha/gamma agonist on Insulin Sensitivity in Patients of Type 2 Diabetes with Hypertriglyceridemia-Randomized double-blind placebo-controlled trial. Sci Rep 2019;9:19017.

42. Rastogi A, Dunbar RL, Thacker HP, et al. Abrogation of postprandial triglyceridemia with dual PPAR alpha/gamma agonist in type 2 diabetes mellitus: a randomized, placebo-controlled study. Acta Diabetol 2020;57:809–818.

43. Kong AP, Yamasaki A, Ozaki R, et al. A randomized-controlled trial to investigate the effects of rivoglitazone, a novel PPAR gamma agonist on glucose-lipid control in type 2 diabetes. Diabetes Obes Metab 2011;13:806–13.

44. Goldstein BJ, Rosenstock J, Anzalone D, et al. Effect of tesaglitazar, a dual PPAR alpha/gamma agonist, on glucose and lipid abnormalities in patients with type 2 diabetes: a 12-week dose-ranging trial. Curr Med Res Opin 2006;22:2575–90.

45. Skrumsager BK, Nielsen KK, Muller M, et al. Ragaglitazar: the pharmacokinetics, pharmacodynamics, and tolerability of a novel dual PPAR alpha and gamma agonist in healthy subjects and patients with type 2 diabetes. J Clin Pharmacol 2003;43:1244–56.

46. Chen W, Fan S, Xie X, et al. Novel PPAR pan agonist, ZBH ameliorates hyperlipidemia and insulin resistance in high fat diet induced hyperlipidemic hamster. PLoS One 2014;9:e96056.

47. Shiomi Y, Yamauchi T, Iwabu M, et al. A Novel Peroxisome Proliferator-activated Receptor (PPAR)alpha Agonist and PPARgamma Antagonist, Z-551, Ameliorates High-fat Diet-induced Obesity and Metabolic Disorders in Mice. J Biol Chem 2015;290:14567–81.

48. Rashid J, Alobaida A, Al-Hilal TA, et al. Repurposing rosiglitazone, a PPAR-gamma agonist and oral antidiabetic, as an inhaled formulation, for the treatment of PAH. J Control Release 2018;280:113–123.

49. Kumar DP, Caffrey R, Marioneaux J, et al. The PPAR alpha/gamma Agonist Saroglitazar Improves Insulin Resistance and Steatohepatitis in a Diet Induced Animal Model of Nonalcoholic Fatty Liver Disease. Sci Rep 2020;10:9330.

50. Gawrieh S, Noureddin M, Loo N, et al. Saroglitazar, a PPAR-alpha/gamma Agonist, for Treatment of Nonalcoholic Fatty Liver Disease: A Randomized Controlled Double-Blind Phase 2 Trial. Hepatology 2021.

51. Chawla A, Barak Y, Nagy L, et al. PPAR-gamma dependent and independent effects on macrophage-gene expression in lipid metabolism and inflammation. Nat Med 2001;7:48–52.

52. Cuzzocrea S, Mazzon E, Di Paola R, et al. The role of the peroxisome proliferator-activated receptor-alpha (PPAR-alpha) in the regulation of acute inflammation. J Leukoc Biol 2006;79:999–1010.

53. Murphy GJ, Holder JC. PPAR-gamma agonists: therapeutic role in diabetes, inflammation and cancer. Trends Pharmacol Sci 2000;21:469–74.

54. Lefebvre P, Chinetti G, Fruchart JC, et al. Sorting out the roles of PPAR alpha in energy metabolism and vascular homeostasis. J Clin Invest 2006;116:571–80.

55. Dubois V, Eeckhoute J, Lefebvre P, et al. Distinct but complementary contributions of PPAR isotypes to energy homeostasis. J Clin Invest 2017;127:1202–1214.

56. Christofides A, Konstantinidou E, Jani C, et al. The role of peroxisome proliferator-activated receptors (PPAR) in immune responses. Metabolism 2021;114:154338.

57. Ju Z, Su M, Hong J, et al. Anti-inflammatory effects of an optimized PPAR-gamma agonist via NF-kappaB pathway inhibition. Bioorg Chem 2020;96:103611.

58. Sugawara K, Olson TS, Moskaluk CA, et al. Linkage to peroxisome proliferator-activated receptor-gamma in SAMP1/YitFc mice and in human Crohn’s disease. Gastroenterology 2005;128:351–60.

59. Kozaiwa K, Sugawara K, Smith MF, Jr., et al. Identification of a quantitative trait locus for ileitis in a spontaneous mouse model of Crohn’s disease: SAMP1/YitFc. Gastroenterology 2003;125:477–90.

60. Dignass A, Marteau P. Mesalamine in the treatment of active Crohn’s disease. Gastroenterology 2005;128:245-6; author reply 246.

61. Hugot JP. PPAR and Crohn’s disease: another piece of the puzzle? Gastroenterology 2005;128:500–3.

62. Yamamoto-Furusho JK, Penaloza-Coronel A, Sanchez-Munoz F, et al. Peroxisome proliferator-activated receptor-gamma (PPAR-gamma) expression is downregulated in patients with active ulcerative colitis. Inflamm Bowel Dis 2011;17:680–1.

63. Carr RM, Patel A, Bownik H, et al. Intestinal Inflammation Does Not Predict Nonalcoholic Fatty Liver Disease Severity in Inflammatory Bowel Disease Patients. Dig Dis Sci 2017;62:1354–1361.

64. Dragasevic S, Stankovic B, Kotur N, et al. Metabolic Syndrome in Inflammatory Bowel Disease: Association with Genetic Markers of Obesity and Inflammation. Metab Syndr Relat Disord 2020;18:31–38.

65. Nagahori M, Hyun SB, Totsuka T, et al. Prevalence of metabolic syndrome is comparable between inflammatory bowel disease patients and the general population. J Gastroenterol 2010;45:1008–13.

66. Kountouras J, Doulberis M, Polyzos SA, et al. Metabolic syndrome components including high abdominal obesity and sarcopenia in patients with inflammatory bowel disease. Ann Gastroenterol 2019;32:214.

67. Verdugo-Meza A, Ye J, Dadlani H, et al. Connecting the Dots Between Inflammatory Bowel Disease and Metabolic Syndrome: A Focus on Gut-Derived Metabolites. Nutrients 2020;12.

68. Michalak A, Mosinska P, Fichna J. Common links between metabolic syndrome and inflammatory bowel disease: Current overview and future perspectives. Pharmacol Rep 2016;68:837–46.

69. Morshedzadeh N, Rahimlou M, Shahrokh S, et al. The effects of flaxseed supplementation on metabolic syndrome parameters, insulin resistance and inflammation in ulcerative colitis patients: An open-labeled randomized controlled trial. Phytother Res 2021.

70. Sourianarayanane A, Garg G, Smith TH, et al. Risk factors of non-alcoholic fatty liver disease in patients with inflammatory bowel disease. J Crohns Colitis 2013;7:e279–85.

71. Magri S, Paduano D, Chicco F, et al. Nonalcoholic fatty liver disease in patients with inflammatory bowel disease: Beyond the natural history. World J Gastroenterol 2019;25:5676–5686.

72. Murray A, Nguyen TM, Parker CE, et al. Oral 5-aminosalicylic acid for induction of remission in ulcerative colitis. Cochrane Database Syst Rev 2020;8:CD000543.

73. Schroeder KW, Tremaine WJ, Ilstrup DM. Coated oral 5-aminosalicylic acid therapy for mildly to moderately active ulcerative colitis. A randomized study. N Engl J Med 1987;317:1625–9.

74. Schreiber S, Howaldt S, Raedler A. Oral 4-aminosalicylic acid versus 5-aminosalicylic acid slow release tablets. Double blind, controlled pilot study in the maintenance treatment of Crohn’s ileocolitis. Gut 1994;35:1081–5.

75. Ham M, Moss AC. Mesalamine in the treatment and maintenance of remission of ulcerative colitis. Expert Rev Clin Pharmacol 2012;5:113–23.

76. Gaggini M, Morelli M, Buzzigoli E, et al. Non-alcoholic fatty liver disease (NAFLD) and its connection with insulin resistance, dyslipidemia, atherosclerosis and coronary heart disease. Nutrients 2013;5:1544–60.

77. Godoy-Matos AF, Silva Junior WS, Valerio CM. NAFLD as a continuum: from obesity to metabolic syndrome and diabetes. Diabetol Metab Syndr 2020;12:60.

78. Targher G, Byrne CD, Tilg H. NAFLD and increased risk of cardiovascular disease: clinical associations, pathophysiological mechanisms and pharmacological implications. Gut 2020;69:1691–1705.

79. Alende-Castro V, Alonso-Sampedro M, Vazquez-Temprano N, et al. Factors influencing erythrocyte sedimentation rate in adults: New evidence for an old test. Medicine (Baltimore) 2019;98:e16816.

80. Pravenec M, Kajiya T, Zidek V, et al. Effects of human C-reactive protein on pathogenesis of features of the metabolic syndrome. Hypertension 2011;57:731–7.

81. Devaraj S, Singh U, Jialal I. Human C-reactive protein and the metabolic syndrome. Curr Opin Lipidol 2009;20:182–9.

82. Ridker PM, Hennekens CH, Buring JE, et al. C-reactive protein and other markers of inflammation in the prediction of cardiovascular disease in women. N Engl J Med 2000;342:836–43.

83. Ridker PM, Buring JE, Cook NR, et al. C-reactive protein, the metabolic syndrome, and risk of incident cardiovascular events: an 8-year follow-up of 14 719 initially healthy American women. Circulation 2003;107:391–7.

84. Luck H, Tsai S, Chung J, et al. Regulation of obesity-related insulin resistance with gut anti-inflammatory agents. Cell Metab 2015;21:527–42.

85. Rabot S, Membrez M, Blancher F, et al. High fat diet drives obesity regardless the composition of gut microbiota in mice. Sci Rep 2016;6:32484.

86. Marungruang N, Fak F, Tareke E. Heat-treated high-fat diet modifies gut microbiota and metabolic markers in apoe-/-mice. Nutr Metab (Lond) 2016;13:22.

87. Serino M, Luche E, Gres S, et al. Metabolic adaptation to a high-fat diet is associated with a change in the gut microbiota. Gut 2012;61:543–53.

88. Shi C, Li H, Qu X, et al. High fat diet exacerbates intestinal barrier dysfunction and changes gut microbiota in intestinal-specific ACF7 knockout mice. Biomed Pharmacother 2019;110:537–545.

89. Hamilton MK, Boudry G, Lemay DG, et al. Changes in intestinal barrier function and gut microbiota in high-fat diet-fed rats are dynamic and region dependent. Am J Physiol Gastrointest Liver Physiol 2015;308:G840–51.

90. Thevaranjan N, Puchta A, Schulz C, et al. Age-Associated Microbial Dysbiosis Promotes Intestinal Permeability, Systemic Inflammation, and Macrophage Dysfunction. Cell Host Microbe 2017;21:455–466 e4.

91. Chen P, Starkel P, Turner JR, et al. Dysbiosis-induced intestinal inflammation activates tumor necrosis factor receptor I and mediates alcoholic liver disease in mice. Hepatology 2015;61:883–94.

92. Butto LF, Haller D. Dysbiosis in intestinal inflammation: Cause or consequence. Int J Med Microbiol 2016;306:302–309.

93. Nagpal R, Newman TM, Wang S, et al. Obesity-Linked Gut Microbiome Dysbiosis Associated with Derangements in Gut Permeability and Intestinal Cellular Homeostasis Independent of Diet. J Diabetes Res 2018;2018:3462092.

94. Porras D, Nistal E, Martinez-Florez S, et al. Intestinal Microbiota Modulation in Obesity-Related Non-alcoholic Fatty Liver Disease. Front Physiol 2018;9:1813.

95. Bray GA. Current status of intestinal bypass surgery in the treatment of obesity. Diabetes 1977;26:1072–9.

96. Debedat J, Clement K, Aron-Wisnewsky J. Gut Microbiota Dysbiosis in Human Obesity: Impact of Bariatric Surgery. Curr Obes Rep 2019;8:229–242.

97. Aron-Wisnewsky J, Prifti E, Belda E, et al. Major microbiota dysbiosis in severe obesity: fate after bariatric surgery. Gut 2019;68:70–82.

98. Sato J, Kanazawa A, Ikeda F, et al. Gut dysbiosis and detection of “live gut bacteria” in blood of Japanese patients with type 2 diabetes. Diabetes Care 2014;37:2343–50.

99. Rosario D, Benfeitas R, Bidkhori G, et al. Understanding the Representative Gut Microbiota Dysbiosis in Metformin-Treated Type 2 Diabetes Patients Using Genome-Scale Metabolic Modeling. Front Physiol 2018;9:775.

100. Noureldein M, Bitar S, Youssef N, et al. Butyrate Modulates Diabetes-Linked Gut Dysbiosis: Epigenetic and Mechanistic Modifications. J Mol Endocrinol 2019.

101. Lee HC, Yu SC, Lo YC, et al. A high linoleic acid diet exacerbates metabolic responses and gut microbiota dysbiosis in obese rats with diabetes mellitus. Food Funct 2019;10:786–798.

102. Holmes D. Gut microbiota: Antidiabetic drug treatment confounds gut dysbiosis associated with type 2 diabetes mellitus. Nat Rev Endocrinol 2016;12:61.

103. Shen F, Zheng RD, Sun XQ, et al. Gut microbiota dysbiosis in patients with non-alcoholic fatty liver disease. Hepatobiliary Pancreat Dis Int 2017;16:375–381.

104. Da Silva HE, Teterina A, Comelli EM, et al. Nonalcoholic fatty liver disease is associated with dysbiosis independent of body mass index and insulin resistance. Sci Rep 2018;8:1466.

105. Chi Y, Lin Y, Lu Y, et al. Gut microbiota dysbiosis correlates with a low-dose PCB126-induced dyslipidemia and non-alcoholic fatty liver disease. Sci Total Environ 2019;653:274–282.

106. Boursier J, Mueller O, Barret M, et al. The severity of nonalcoholic fatty liver disease is associated with gut dysbiosis and shift in the metabolic function of the gut microbiota. Hepatology 2016;63:764–75.

107. Bertin B, Dubuquoy L, Colombel JF, et al. PPAR-gamma in ulcerative colitis: a novel target for intervention. Curr Drug Targets 2013;14:1501–7.

108. Rousseaux C, El-Jamal N, Fumery M, et al. The 5-aminosalicylic acid antineoplastic effect in the intestine is mediated by PPARgamma. Carcinogenesis 2013;34:2580–6.

109. Wang Z, Koonen D, Hofker M, et al. 5-aminosalicylic acid improves lipid profile in mice fed a high-fat cholesterol diet through its dual effects on intestinal PPARgamma and PPARalpha. PLoS One 2018;13:e0191485.

110. Bazzi C, Sinico RA, Petrini C, et al. Low doses of drugs able to alter intestinal mucosal permeability to food antigens (5-aminosalicylic acid and sodium cromoglycate) do not reduce proteinuria in patients with IgA nephropathy: a preliminary noncontrolled trial. Nephron 1992;61:192–5.

111. Di Paolo MC, Merrett MN, Crotty B, et al. 5-Aminosalicylic acid inhibits the impaired epithelial barrier function induced by gamma interferon. Gut 1996;38:115–9.

112. Cannon AR, Akhtar S, Hammer AM, et al. Effects of Mesalamine Treatment on Gut Barrier Integrity After Burn Injury. J Burn Care Res 2016;37:283–92.

113. Xu J, Chen N, Wu Z, et al. 5-Aminosalicylic Acid Alters the Gut Bacterial Microbiota in Patients With Ulcerative Colitis. Front Microbiol 2018;9:1274.

114. Andrews CN, Griffiths TA, Kaufman J, et al. Mesalazine (5-aminosalicylic acid) alters faecal bacterial profiles, but not mucosal proteolytic activity in diarrhoea-predominant irritable bowel syndrome. Aliment Pharmacol Ther 2011;34:374–83.

115. Xue L, Huang Z, Zhou X, et al. The possible effects of mesalazine on the intestinal microbiota. Aliment Pharmacol Ther 2012;36:813–4.

116. Jun X, Ning C, Yang S, et al. Alteration of Fungal Microbiota After 5-ASA Treatment in UC Patients. Inflamm Bowel Dis 2019.

117. Kaiser GC, Yan F, Polk DB. Mesalamine blocks tumor necrosis factor growth inhibition and nuclear factor kappaB activation in mouse colonocytes. Gastroenterology 1999;116:602–9.

118. Bantel H, Berg C, Vieth M, et al. Mesalazine inhibits activation of transcription factor NF-kappaB in inflamed mucosa of patients with ulcerative colitis. Am J Gastroenterol 2000;95:3452–7.

119. Qu T, Wang E, Jin B, et al. 5-Aminosalicylic acid inhibits inflammatory responses by suppressing JNK and p38 activity in murine macrophages. Immunopharmacol Immunotoxicol 2017;39:45–53.

120. Stolfi C, Fina D, Caruso R, et al. Cyclooxygenase-2-dependent and -independent inhibition of proliferation of colon cancer cells by 5-aminosalicylic acid. Biochem Pharmacol 2008;75:668–76.

121. Sharon P, Ligumsky M, Rachmilewitz D, et al. Role of prostaglandins in ulcerative colitis. Enhanced production during active disease and inhibition by sulfasalazine. Gastroenterology 1978;75:638–40.

122. Collier HO, Francis AA, McDonald-Gibson WJ, et al. Inhibition of prostaglandin biosynthesis by sulphasalazine and its metabolites. Prostaglandins 1976;11:219–25.

123. Egan LJ, Mays DC, Huntoon CJ, et al. Inhibition of interleukin-1-stimulated NF-kappaB RelA/p65 phosphorylation by mesalamine is accompanied by decreased transcriptional activity. J Biol Chem 1999;274:26448–53.

124. Allgayer H, Kruis W. Aminosalicylates: potential antineoplastic actions in colon cancer prevention. Scand J Gastroenterol 2002;37:125–31.

125. Bus PJ, Nagtegaal ID, Verspaget HW, et al. Mesalazine-induced apoptosis of colorectal cancer: on the verge of a new chemopreventive era? Aliment Pharmacol Ther 1999;13:1397–402.

126. Reinacher-Schick A, Seidensticker F, Petrasch S, et al. Mesalazine changes apoptosis and proliferation in normal mucosa of patients with sporadic polyps of the large bowel. Endoscopy 2000;32:245–54.

127. Olaisen M, Spigset O, Flatberg A, et al. Mucosal 5-aminosalicylic acid concentration, drug formulation and mucosal microbiome in patients with quiescent ulcerative colitis. Aliment Pharmacol Ther 2019;49:1301–1313.

128. Dahl JU, Gray MJ, Bazopoulou D, et al. The anti-inflammatory drug mesalamine targets bacterial polyphosphate accumulation. Nat Microbiol 2017;2:16267.

129. Dai L, Tang Y, Zhou W, et al. Gut Microbiota and Related Metabolites Were Disturbed in Ulcerative Colitis and Partly Restored After Mesalamine Treatment. Front Pharmacol 2020;11:620724.

